# OraSure InteliSwab^®^ Rapid Antigen Test performance with the SARS-CoV-2 Variants of Concern Alpha, Beta, Gamma, Delta, and Omicron

**DOI:** 10.1101/2022.02.02.22270254

**Authors:** Zachary A. Weishampel, Janean Young, Mark Fischl, Robert J. Fischer, Irene Owusu Donkor, Jade C. Riopelle, Jonathan E. Schulz, Julia R. Port, Taylor A. Saturday, Neeltje van Doremalen, Jody D. Berry, Vincent J. Munster, Claude Kwe Yinda

## Abstract

The emergence of SARS-CoV-2 in the human population and the resulting COVID-19 pandemic has led to the development of various diagnostic tests. The OraSure InteliSwab^®^ COVID-19 Rapid Test is a recently developed and FDA emergency use authorized rapid antigen-detecting test that functions as a lateral flow device targeting the nucleocapsid protein. Due to SARS-CoV-2 evolution, there is a need to evaluate the sensitivity of rapid antigen-detecting tests for new variants, especially variants of concern like Omicron. In this study, the sensitivity of the OraSure InteliSwab^®^ Test was investigated using cultured strains of the known variants of concern (VOCs, Alpha, Beta, Gamma, Delta, and Omicron) and the ancestral lineage (lineage A). Based on dilution series in cell culture medium, an approximate limit of detection for each variant was determined. The OraSure InteliSwab^®^ Test showed an overall comparable performance using recombinant nucleocapsid protein and different cultured variants with recorded limits of detection ranging between 3.77 × 10^5^ and 9.13 × 10^5^ RNA copies/mL. Finally, the sensitivity was evaluated using oropharyngeal swabs from Syrian golden hamsters inoculated with the 6 VOCs. Ultimately, the OraSure InteliSwab^®^ COVID-19 Rapid Test showed no decrease in sensitivity between the ancestral SARS-CoV-2 strain and any VOCs including Omicron.

## 1. Introduction

Since the emergence of SARS-CoV-2, a wide variety of diagnostic assays have been developed. These assays primarily use quantitative real-time reverse transcription polymerase chain reactions (qRT-PCRs) which detect viral RNA. Due to their high sensitivity and specificity, qRT-PCRs function as the gold standard for COVID-19 diagnostics [1]. However, qRT-PCR-based diagnostics require advanced laboratory infrastructure and trained personnel. Furthermore, the relatively long time to receive results could hamper direct decision making.

Another diagnostic assay is the rapid antigen-detecting test (RDT). RDTs can be done at home, produce results within hours [2-5], and complement qRT-PCR-based diagnostics [6]. As the name suggests, RDTs are based on the detection of antigen presence.

The emergence of new variants of concern (VOCs) highlights the need to continuously validate current diagnostic assays. Here, we assess the sensitivity of the OraSure InteliSwab^®^ using all currently identified VOCs (Alpha, Beta, Gamma, Delta, and Omicron [7]) compared to the ancestral lineage (lineage A) of SARS-CoV-2. In addition, we employ the Syrian hamster COVID-19 model to determine the InteliSwab^®^ performance in a controlled infection environment.

## 2. Materials and Methods

### 2.1 Recombinant Nucleocapsid Protein Assay

SARS-CoV-2 nucleocapsid proteins (NPs) of described variants were expressed with an N-terminus poly-histidine tag in BL21(DE3) cells. Affinity capture of the protein was performed using nickel sepharose HP, followed by a buffer exchange into Tris/NaCl using a GE HiTrap 26/10 desalting column. The lineage A variant was expressed using reference sequence QHD43423.2, amino acids 1-419. Subsequent SARS-CoV-2 variant mutations were cloned via mutagenesis and sequenced confirmed. All recombinant NPs used for device testing were > 95% purity. Expression levels of purified NP were 80-150 mg/L depending on the variant. To load sample on the test device, 50 µL of sample was introduced at the center of the flat pad. With the developer solution vials resting in the test stand’s slots, the loaded test devices were placed in separate vials containing the developer solution. Each test device was left in the developer solution for 30 minutes after which the result was read. A positive result was a red line at the test and control zones (Figure 1a).

**Figure 1.**
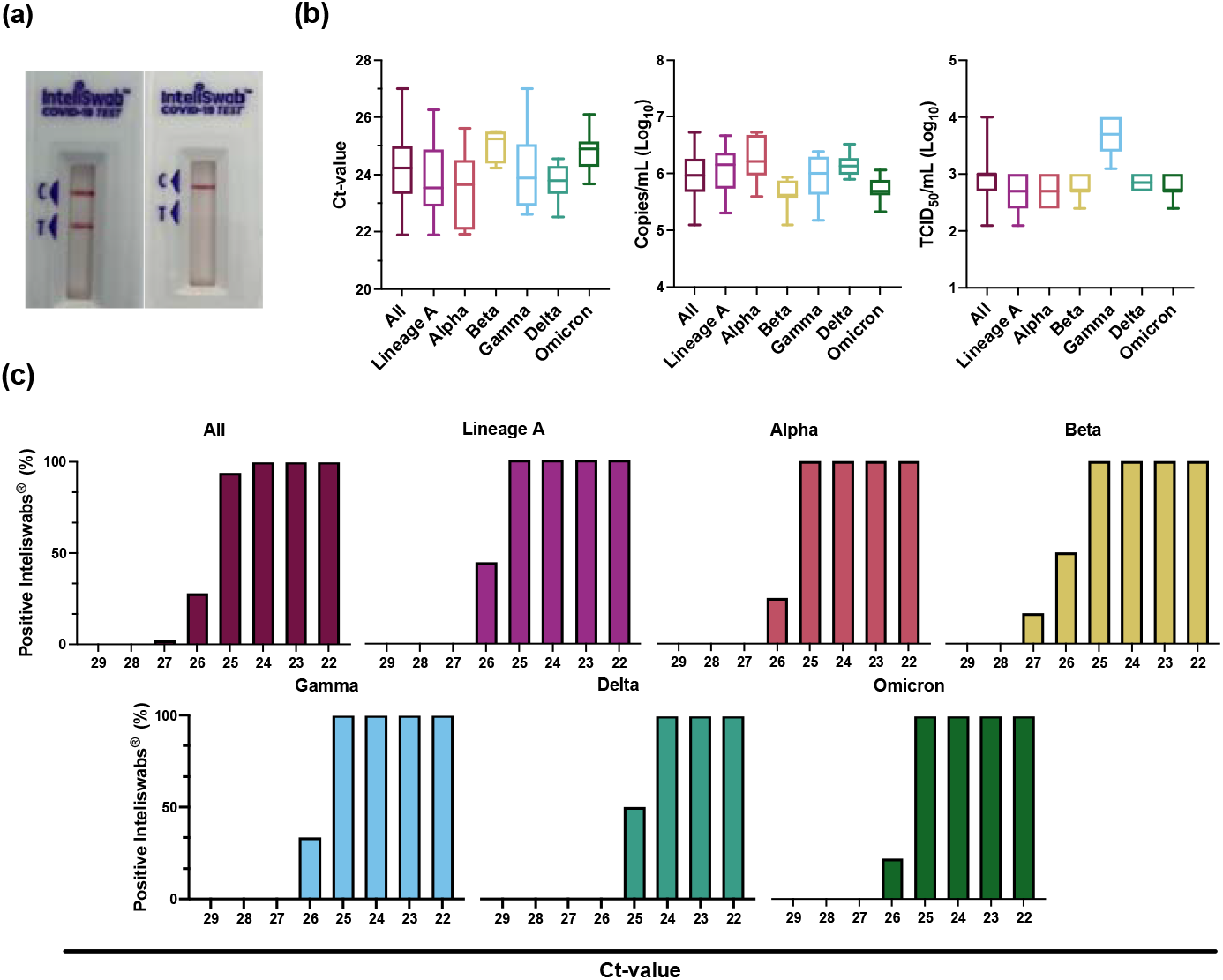
InteliSwab^®^ Test detection ability for SARS-CoV-2 variants. Cell culture stocks of the variants were serially diluted, and viral load was measured by gRNA qRT-PCR. (**a**) Comparison of a positive InteliSwab^®^ Test (left) and a negative InteliSwab^®^ Test (right). **(b**) Box plots displaying the distribution of positive InteliSwab^®^ Tests across the 6 variants compared to the Ct-values, RNA copies/mL, and TCID50/mL. The positive InteliSwabs^®^ are from the 2-fold dilution series for each variant (from left to right, N = 140, 31, 27, 13, 31, 18, and 20). Shown is the median, minimum, and maximum. (**c**) Bar charts representing the percentage of positive InteliSwab^®^ Tests compared to Ct-values. Initial Ct-values were rounded to the nearest whole number.

### 2.2 Cells and viruses

The SARS-CoV-2 isolates used in this study are summarized in Table S1. Virus propagation was performed in VeroE6 cells in DMEM supplemented with 2% fetal bovine serum, 1 mM L-glutamine, 50 U/mL penicillin and 50 µg/mL streptomycin (DMEM2). VeroE6 cells were maintained in DMEM supplemented with 10% fetal bovine serum, 1 mM L-glutamine, 50 U/mL penicillin and 50 µg/mL streptomycin. At regular intervals, mycoplasma testing was performed. No mycoplasma and no contaminants were detected. Sequencing from viral stocks included libraries prepared from Stranded Total RNA Prep Ligation with Ribo-Zero Plus kit per manufacturer’s protocol (Illumina) and sequenced on an Illumina MiSeq at 2 × 150 base pair reads. No nucleotide change was found > 5%.

### 2.3 InteliSwab^®^ assay on SARS-CoV-2 isolates

Prior to dilutions and tests, each SARS-CoV-2 stock variant was inactivated using irradiation with 2 Mrad. To evaluate the effect of the irradiation process on the InteliSwab^®^ results, the test was performed on irradiated and unirradiated 10-fold serially diluted Delta variant stocks. The results showed that the sensitivity of OraSure InteliSwab^®^ COVID-19 Rapid Test was the same for both the irradiated and the unirradiated stocks (Figure S1).

Then, 10-fold serial dilutions of irradiated SARS-CoV-2 variants were performed with 1x Dulbecco’s phosphate-buffered saline (PBS). Three OraSure InteliSwab^®^ COVID-19 Rapid Tests were used to test each dilution, making a total of 15 test devices per variant. The test procedure was carried out as above. More precise limits of detection (LODs) for the test were determined by performing 2-fold serial dilutions in 3 replicates for each variant beginning at the 10-fold dilution LOD. Three test devices were used for each dilution. The same procedures for loading samples and analyzing test results were followed for the 10-fold serial dilutions. The final LOD for each variant was determined to be the minimum concentration (dilution) for which all 9 test devices were positive.

### 2.4 Ethics Statement

Animal experiments were conducted in an AAALAC International-accredited facility and approved by the Rocky Mountain Laboratories Institutional Care and Use Committee following the guidelines put forth in the Guide for the Care and Use of Laboratory Animals 8th edition, the Animal Welfare Act, United States Department of Agriculture and the United States Public Health Service Policy on the Humane Care and Use of Laboratory Animals.

The Institutional Biosafety Committee (IBC) approved work with SARS-CoV-2 strains under BSL3 conditions. Virus inactivation of all samples was performed according to IBC-approved standard operating procedures for the removal of specimens from high containment areas.

### 2.5 Animal Experiment

Four-six-week-old Syrian golden hamsters (N = 6 per group, Envigo Indianapolis) were challenged intranasally with 40 µL containing 1 × 10^3^ Median Tissue Culture Infectious Dose (TCID50)/mL virus in sterile DMEM. All virus stocks were full genome sequenced, no SNPs > 10% in spike or nucleoprotein were detected. Weights were recorded daily. Oropharyngeal swabs were collected in 1 mL of DMEM2 on day post infection (DPI) 1-7. Fifty µL of media was pipetted onto the absorbent pad of the InteliSwab^®^ as outlined in the instructions for use, described previously.

### 2.6 RNA Extraction and qRT-PCR

RNA was extracted from 140 µL of sample for each dilution with the Qiagen QIAamp Viral RNA Kit according to the manufacturer’s instructions with an elution volume of 60 µL. Following the extraction, copies of genomic RNA was determined by qRT-PCR with the TaqMan™ Fast Virus One-Step Master Mix and QuantStudio 6 Flex Real-Time PCR System from Applied Biosystems according to manufacturer’s instructions. Ten-fold dilutions of SARS-CoV-2 standards with known copy numbers were used to construct a standard curve and calculate copies/mL.

## 3. Results

Through the use of monoclonal antibodies, the OraSure InteliSwab^®^ COVID-19 Rapid Test detects the conserved SARS-CoV-2 nucleocapsid protein (NP). To assess the performance of the InteliSwab^®^ for different VOCs, we first generated recombinant NP for the identified SARS-CoV-2 VOCs and the ancestral lineage A (Table 1). Sample dilutions were prepared in phosphate buffered saline (PBS) and 50 µL of the dilution was used on each RDT. In order to determine the limit of detection (LOD), the assay was performed for each of the variant NPs with N = 20 tests. Limited variation was observed between the different NPs; the NP LOD was either 0.313 ng/mL for Alpha and Gamma or 0.469 ng/mL for lineage A, Beta, Delta, and Omicron (Table 2).

**Table 1.**
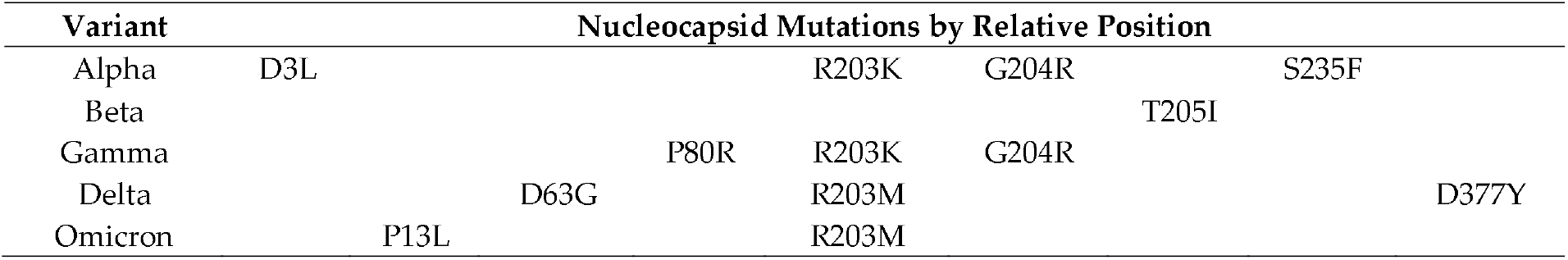
Nucleocapsid protein mutations of the SARS-CoV-2 variants

**Table 2.**
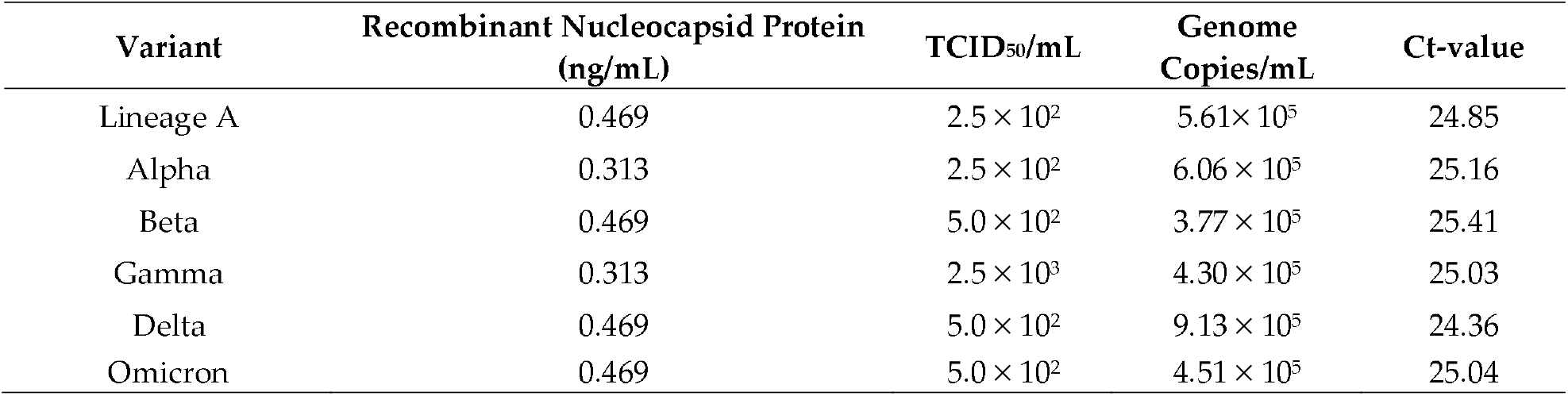
InteliSwab^®^ limit of detections for SARS-CoV-2 variants

| Variant | Recombinant Nucleocapsid Protein (ng/mL) | TCID<sub>50</sub>/mL | Genome Copies/mL | Ct-value |
| --- | --- | --- | --- | --- |
| Lineage A | 0.469 | $2.5 \times 10^2$ | $5.61 \times 10^5$ | 24.85 |
| Alpha | 0.313 | $2.5 \times 10^2$ | $6.06 \times 10^5$ | 25.16 |
| Beta | 0.469 | $5.0 \times 10^2$ | $3.77 \times 10^5$ | 25.41 |
| Gamma | 0.313 | $2.5 \times 10^3$ | $4.30 \times 10^5$ | 25.03 |
| Delta | 0.469 | $5.0 \times 10^2$ | $9.13 \times 10^5$ | 24.36 |
| Omicron | 0.469 | $5.0 \times 10^2$ | $4.51 \times 10^5$ | 25.04 |

The sensitivity of the RDT was also evaluated using SARS-CoV-2 virus isolates. Virus stocks were serially diluted 10-fold, starting at 1 × 10^5^ median tissue culture infectious dose (TCID50)/mL, and SARS-CoV-2 genome copy number and cycle-threshold value (Ct-value) were determined. Next, 50 µL of each dilution was tested in triplicate. The lowest virus concentration at which all 3 RDTs were positive was 1 × 10^3^ TCID50/mL for all variants except Gamma, which was 1 × 10^4^ TCID50/mL. These lowest virus dilutions with 3 positive RDTs represented Ct-values between 21 and 23 and corresponded to genome copy numbers between 7.72 × 10^6^ and 1.59 × 10^6^ copies/mL.

To differentiate further, 2-fold dilutions were performed in triplicate, beginning at the lowest virus concentration with 3 positive RDTs from the 10-fold dilutions described above. Each of these dilutions was then evaluated in triplicate using 50 µL on the RDT. The LOD was defined as the lowest virus concentration with all 9 positive RDTs. The LODs based on TCID50/mL were 2.50 × 10^2^ for both lineage A and Alpha, 5.00 × 10^2^ for Beta, Delta, and Omicron, and 2.50 × 10^3^ for Gamma (Table 2, Figure 1b). For Ct-values, the LODs were 24.85 for lineage A, 25.16 for Alpha, 25.41 for Beta, 25.03 for Gamma, 24.36 for Delta, and 25.04 for Omicron (Figure 1c, Figure S2). These values corresponded to genome copies/mL of 5.61 × 10^5^, 6.06 × 10^5^, 3.77 × 10^5^, 4.30 × 10^5^, 9.13 × 10^5^, and 4.51 × 10^5^, respectively.

To determine the performance of the RDT in a controlled infection environment, we used the SARS-CoV-2 Syrian hamster model. Six 4-6 week old male and female Syrian golden hamsters were intranasally inoculated with 1 × 10^3^ TCID50/mL for each variant. Oropharyngeal swabs were collected daily in 1 mL DMEM containing 2% FBS, 2 mM L-glutamine, and 100 units/mL penicillin/streptomycin. Swabs were analyzed by qRT-PCR and tested on the InteliSwab^®^ (1 RDT per hamster per day, 50 µL). On average, all 6 animals were positive for 4 consecutive days, starting at 1 day post infection (DPI), for lineage A, Alpha, Beta, Gamma, and Delta. For Omicron, however, the RDTs were positive 6/6 on 2 DPI, 5/6 on 3 DPI, and 3/6 on 3 and 4 DPI. At 6 and 7 DPI, the tests were all negative (Figure 2a). This was explained by decreased shedding after Omicron infection as compared to the other variants, and not by decreased test sensitivity (Figure 2b). The overall Ct-values of positive RDTs were comparable between the different viruses and were at most 24 – 26 (Figure 2c). Combined with the reduction in shedding in this animal model, this suggests that the functioning of the RDT is directly related to the infection kinetics within the host.

**Figure 2.**
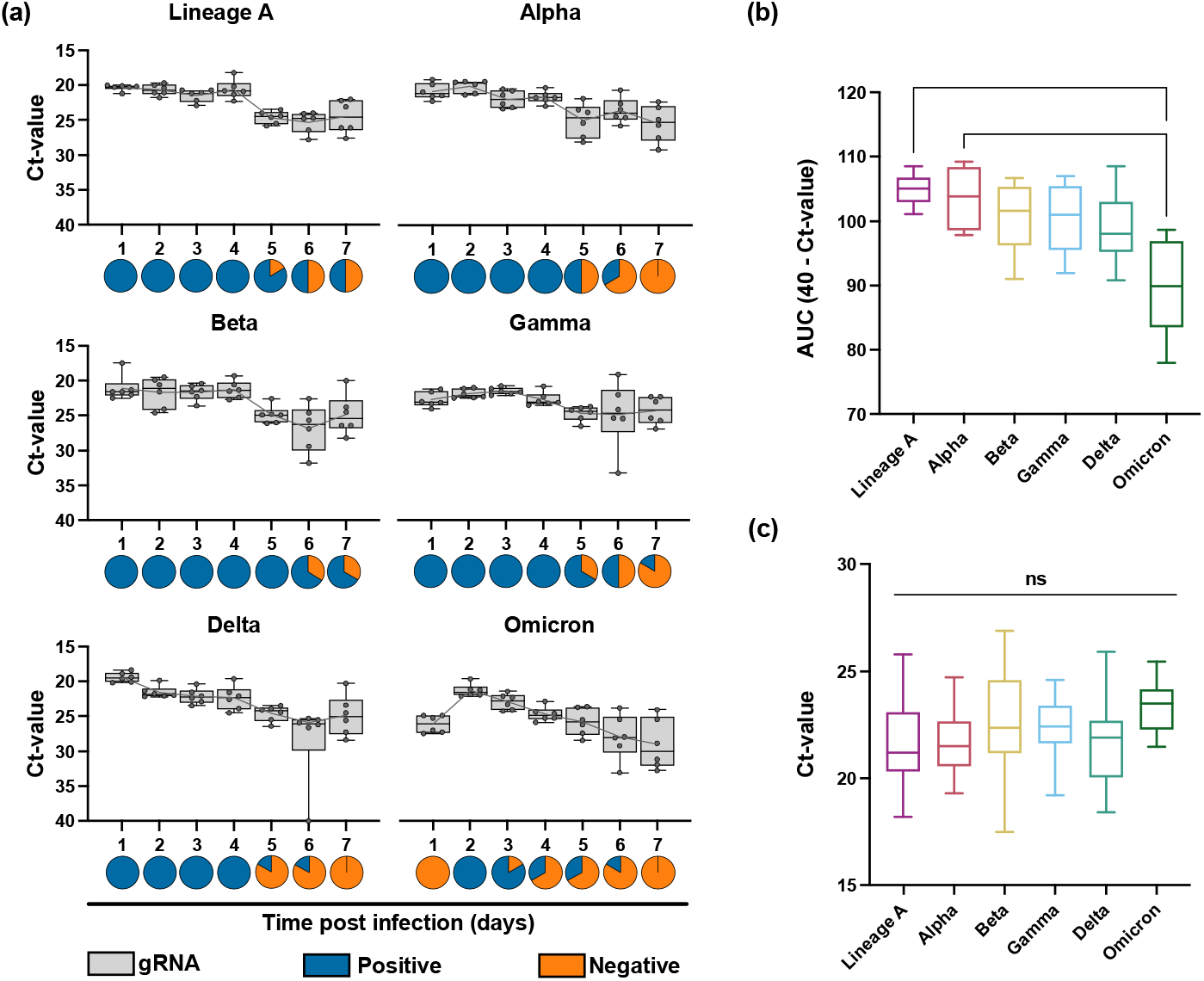
InteliSwab^®^ results for SARS-CoV-2 variants in Syrian golden hamsters. For each variant, animals (N = 6) were inoculated with 1 × 10^3^ TCID50/mL. Oropharyngeal swabs were taken for 7 days. **(a)** Box plots depicting viral load (gRNA) in swabs across the 7 days; Box and whiskers represent the median, minimum, maximum, and individual Ct-values. Gray lines represent the mean Ct-value of oropharyngeal swabs for each day. Pie charts show results of oropharyngeal swabs tested by InteliSwab^®^; blue = positive, orange = negative. For b and c, Statistical significance was measured by the Kruskal-Wallis test. ns = not significant, *P < 0.05, **P < 0.001. **(b)** Box plots showing cumulative (area under the curve (AUC) analysis) respiratory shedding of SARS-CoV-2 variants viral load in oropharyngeal swabs measured by gRNA. Box and whiskers represent the median, minimum, and maximum cumulative respiratory shedding. For lineage A and Omicron, P = 0.0083. For Alpha and Omicron, P = 0.0385. **(c**) Box plots representing the positive OraSure InteliSwabs^®^ from all oropharyngeal swabs measured across the 7 days. Box and whiskers represent the median, minimum, and maximum Ct-values (from left to right, N = 35, 28, 38, 32, 26, and 10).

## 4. Discussion

As SARS-CoV-2 continues to evolve in the human population, there is a concern that diagnostic assays originally designed using the ancestral lineage A strain will not be able to detect new variants. Previous studies have demonstrated no variation in sensitivity for some of the RDT tests between the variants Alpha, Beta, Gamma, Delta, and the ancestral strain [8-11]. However only a few studies have included Omicron [12]. Because such a pan-variant of concern antigen test is crucial for public health, there is a need to evaluate RDTs on the current VOCs.

Using recombinant protein and cell-cultured SARS-CoV-2 variants, we observed only minor differences in the sensitivity of the InteliSwab^®^ for Alpha, Beta, Gamma, Delta, and Omicron compared to the initial lineage A variant. Thus, we conclude that the mutations present in the NPs for the current VOCs do not affect the detection ability of this RDT. In addition, based on Ct-values, no difference in sensitivity was observed when using the InteliSwab^®^ on Syrian golden hamsters experimentally infected with the different VOCs. Moreover, the InteliSwab^®^ RDT here demonstrated similar sensitivity to VOCs compared to four other RDTs, which were previously found to have a general LOD between 2 × 10^6^ - 5 × 10^6^ genome copies/mL for lineage A, Alpha, Beta, Gamma, and Delta [8].

Based on our results, recombinant NP can act as a valuable analytical tool for rapid screening the function of RDTs. Differences in post-translational modification in E. coli expressed NP appear not to impact monoclonal antibodies cross recognition of the de facto proteins clinically. This suggests that recombinant NP could be used as a surrogate to live viral testing and would enable easy routine monitoring of emerging VOCs.

It is worth noting that peak shedding of lineage A, Alpha, Beta, Gamma and Delta variants, as determined by the lowest Ct-value, was observed at 1 DPI, while peak shedding of Omicron was observed at 2 DPI. In addition, shedding with Omicron in the hamster model trailed off faster as compared to the other VOCs. Therefore, the delay in the detection ability of this RDT in the samples obtained from Syrian golden hamsters inoculated with Omicron suggests that the overall temporal dynamics in viral shedding might have an impact on the RDT’s ability to detect infection. Preliminary human data suggest a shift towards a delay in the peak of viral shedding in humans with Omicron, similar to what is shown here in the hamster model [13, 14]. This shedding delay could result in an inability for the RDTs to detect a SARS-CoV-2 infection in its early stages.

The OraSure InteliSwab^®^ Test showed comparable sensitivity between the ancestral SARS-CoV-2 strain and all the VOCs including Omicron. Our data would suggest that the InteliSwab^®^ Test is not affected by the current mutations in the NP; however, it is not clear if future mutations may affect the RDT’s sensitivity. Therefore, there is a need to continuously evaluate existing diagnostic tests. With the potential changes in the temporal dynamics of viral shedding in humans, this should also include the real-world performance of RDTs.

## Data Availability

All data produced in the present study are available upon reasonable request to the authors

## Author Contributions

Conceptualization, J.D.B. and V.J.M.; methodology, N.v.D., J.D.B., V.J.M., and C.K.Y.; formal analysis, Z.A.W., J.C.R., T.A.Y., and C.K.Y.; investigation, Z.A.W., J.Y., M.F., R.J.F., I.O.D., J.E.S., J.R.P., N.v.D., J.D.B., V.J.M, and C.K.Y; writing—original draft preparation, Z.A.W. and C.K.Y.; writing—review and editing, all authors; visualization, Z.A.W., J.C.R., T.A.Y., and C.K.Y; supervision, J.D.B., V.J.M., and C.K.Y.; funding acquisition, J.D.B. and V.J.M. All authors have read and agreed to the published version of the manuscript.

## Institutional Review Board Statement

Animal experiments were conducted in an AAALAC International-accredited facility and were approved by the Rocky Mountain Laboratories Institutional Care and Use Committee following the guidelines put forth in the Guide for the Care and Use of Laboratory Animals 8th edition, the Animal Welfare Act, United States Department of Agriculture and the United States Public Health Service Policy on the Humane Care and Use of Laboratory Animals. The Institutional Biosafety Committee (IBC) approved work with infectious SARS-CoV-2 virus strains under BSL3 conditions. Virus inactivation of all samples was performed according to IBC-approved standard operating procedures for the removal of specimens from high containment areas.

## Acknowledgments

We would like to thank Myndi Holbrook, Emmie de Wit, Brandi Williamson, Meaghan Flagg, Kyle Rosenke, Matthew Lewis, Katie Willebrand, Natalie Thornburg, Bin Zhou, Sue Tong, Sujatha Rashid, Ranjan Mukul, Kimberly Stemple, Craig Martens, Kent Barbian, Stacey Ricklefs, Sarah Anzick, Keith Kardos for critical review and the animal care takers for their assistance during the study. The following reagent was obtained through: CDC, hu/USA/CA_CDC_5574/2020, WA1, BEI Resources, NIAID, NIH: SARS-Related Coronavirus 2, Isolate hCoV-19/England/204820464/2020, NR282 54000, contributed by CDC. Severe Acute Respiratory Syndrome-Related Coronavirus 2, Isolate hCoV-19/England/204820464/20200, NR-54000, contributed by Bassam Hallis. B.1.351 (beta variant) Isolate Name: hCoV-19/USA/MD-HP01542/2021 contributed by John Hopkins Bloomberg School of Public Health: Andrew Pekosz. Omicron Isolate name hCoV-19/USA/WI-WSLH-221686/2021 contributed by University of Wisconsin-Madison: Pete Halfman and Yoshihiro Kawaoka.

## Supplementary Materials

**Table S1:** SARS-CoV-2 isolates used in this study

| Virus | WHO Description | PANGO Lineage | GiSAID/Genbank Acc |
| --- | --- | --- | --- |
| SARS-CoV-2/human/USA/WA-CDC-WA1/2020 | Lineage A | WA1 | MN985325 |
| England/204820464/2020 | Alpha | B.1.1.7 | EPI_ISL_683466 |
| USA/MD-HP01542/2021 | Beta | B.1.351 | EPI_ISL_890360 |
| hCoV-19/USA/GA-EHC-2811C/2021 | Gamma | P1 | EPI_ISL_7171744 |
| hCoV-19/USA/KY-CDC-2-4242084/2021 | Delta | B.1.617.2 | EPI_ISL_1823618 |
| hCoV-19/USA/WI-WSLH-221686/2021 | Omicron | B.1.1529 | EPI_ISL_7263803 |

**Figure A1..**
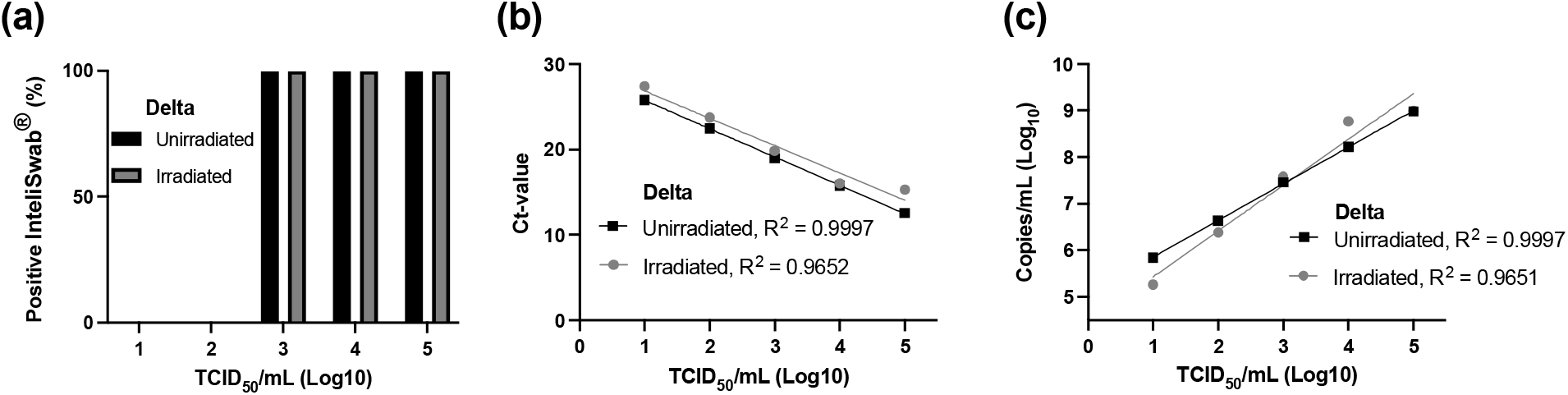
Evaluation of the effects of irradiation on OraSure InteliSwab^®^ results. Two 10-fold dilutions, beginning at 1.00 × 10^5^ TCID50/mL, were performed with live Delta and irradiated Delta. For each dilution, 50 µL was applied to 3 separate OraSure InteliSwab^®^. Irradiation dosage was 2 Mrad. **(a)** Bar plots showing percentage positive OraSure InteliSwab^®^ results for live versus irradiated Delta variant 10-fold dilutions. **(b)** Scatter plot depicting Ct-values for live versus irradiated Delta variant 10-fold dilutions. **(c)** Scatter plot showing RNA copy concentration for live versus irradiated Delta variant 10-fold dilutions.

**Figure S2.**
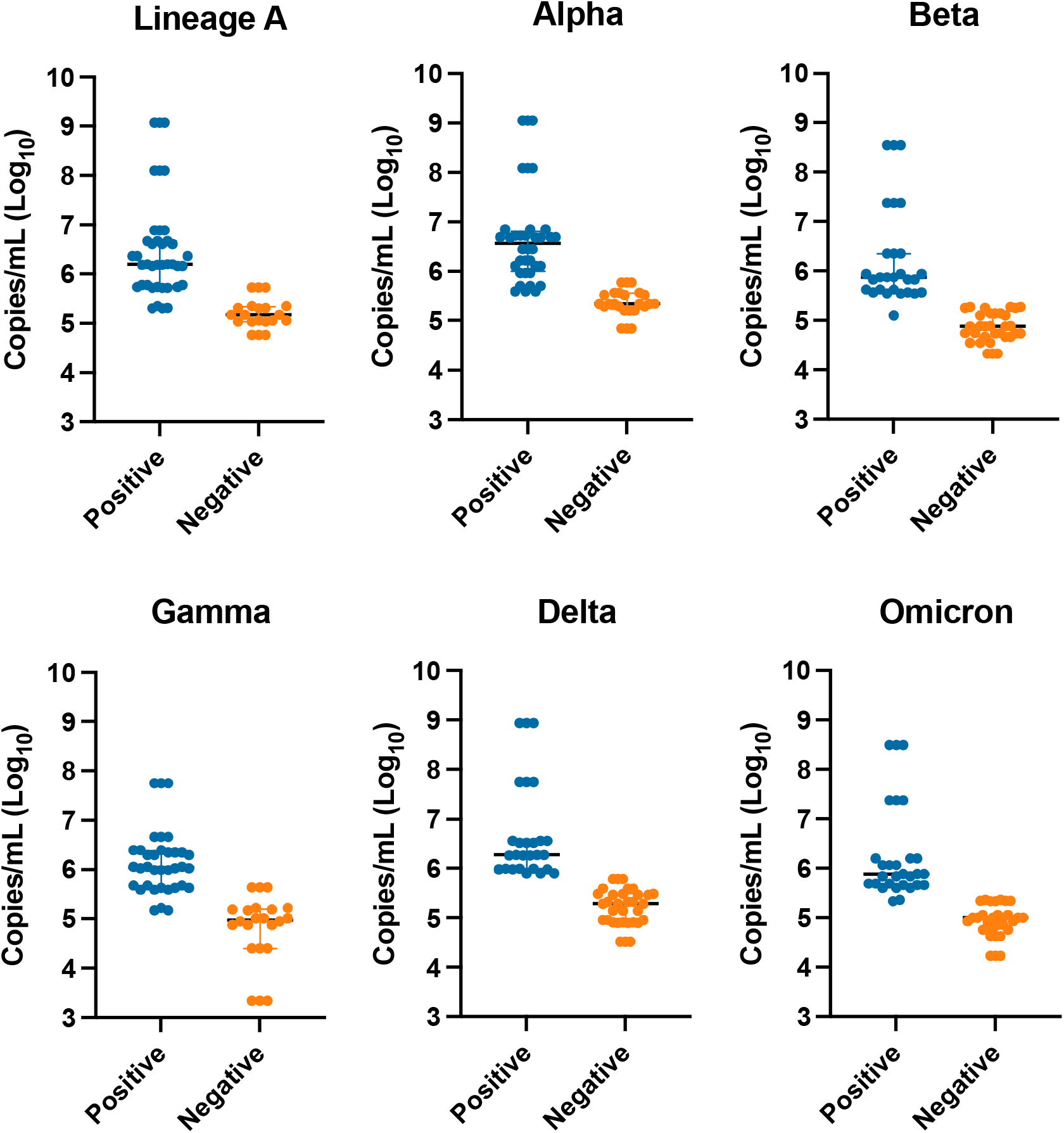
Distribution of all OraSure InteliSwab^®^ results tested on irradiated sample compared to the RNA copies/mL. Scatter plots represent all OraSure InteliSwab^®^ results on irradiated 10-fold and 2-fold dilutions for each variant tested, including lineage A, Alpha, Beta, Gamma, Delta, and Omicron. The blue and orange lines represent the interquartile range for each positive or negative group. The black line represents median.

